# Prevalence and Associated Factors of Female Genital Schistosomiasis in Nkhotakota District, Malawi

**DOI:** 10.1101/2025.10.23.25338636

**Authors:** Dzinkambani Moffat Kambalame, Alex Thawani, Jim Mtambo, Moses Kamzati, Clara Sambani, Faith Mtoso, Tadala Mwale, Jacob Kafulafula, Chifundo Manjawira, Micheal Luhanga, Stellah Lungu, Selemani Ngwira, Chikondi Chimbatata, Memory Moque Ngwira, Bernard Mvula, Billy Nyambalo, Bessie Phiri, Michael Kayange, Matthew Kagoli, Edward Mberu Kamau, Evelyn Chitsa-Banda

**Author notes:** **Corresponding author:** Dzinkambani Moffat Kambalame,.

## Abstract

**Background:** Female Genital Schistosomiasis (FGS) is an critical complication of schistosome infection, especially in schistosomiasis-endemic areas. It is a neglected disease and remains under-recognized in many areas. The prevalence of visual FGS in the endemic areas of Southern Malawi is 26.9%. This is higher in other African countries including Zambia and Tanzania and nosuch studies have been conducted in endemic areas in the Central Region of Malawi. The study aimed to determine the prevalence and risk factors for FGS in Nkhotakota district, Central Malawi.

**Methods:** The cross-sectional study was conducted from November 2024 to March 2025. Two hundred and sixty women (N=260) of childbearing age accessing healthcare services were consecutively sampled from the selected healthcare facilities. An interviewer-guided questionnaire was used to collect data regarding the demographics and FGS symptoms of the participants. Colposcopy was conducted by an experienced gynecological clinical officer to identify FGS lesions and was supervised by a specialist gynecologist. Data were coded, cleaned, and analyzed using the R software (version 4.3.1).

**Results:** The prevalence of FGS using colposcope as a diagnostic tool in women of childbearing age was 32.6%(84/258). Age and a history of cervical cancer screening were associated with increased odds of FGS. The odds of FGS increased by 5% for each additional year (OR, 1.05; 95% CI: 1.02–1.09; p = 0.003). Women with a history of cervical cancer screening were twice as likely to have FGS than those without a history of cervical cancer screening (OR, 1.91; 95% CI: 1.01–3.70; p = 0.05).

**Conclusion:** We found a huge burden of undetected FGS in Nkhotakota district, Central Malawi. Integrated interventions are needed to reduce the burden of FGS and improve its early detection, particularly in schistomiasis-endemic areas.

## 1 Background

Female Genital Schistosomiasis (FGS) is a disease caused by the parasite *Schistosoma haematobium* that contracts in contaminated freshwater(1,2). This condition affects the female reproductive tract and leads to a range of gynecological manifestations. Common complications include infertility, spontaneous abortion, ectopic pregnancy, an increased risk of HIV infection, genital ulcers, premature birth, and low birth weight(1,3). There is evidence that this condition remains neglected in most national healthcare systems as it is given low priority in terms of diagnosis, treatment, and other public health interventions(4,5). This neglect may result in poor reproductive health outcomes among women owing to a lack of adequate support in terms of prevention, diagnosis, and treatment strategies.

FGS is more prevalent in sub-Saharan Africa (SSA). As of 2023, it is estimated that approximately 56 million women and girls will be affected in the region(6). Sub-Saharan Africa accounts for up to 90% of the global FGS cases(7). The FGS is estimated to contribute to approximately 280,000 deaths annually in the region(7). Considering this prevalence, there is a need to understand the factors associated with the burden of FGS to develop and implement appropriate strategies for addressing this challenge.

Furthermore, diagnosis of FGS remains a challenge in SSA, affecting its detection, treatment, and other appropriate healthcare interventions. Poor diagnosis is attributed to several factors, such as limited awareness among healthcare providers, lack of access to diagnostic tools, and overlap of FGS symptoms with other reproductive tract infections and conditions(1,8). This contributes to underreporting and mismanagement of the condition and poor treatment modalities, thereby increasing the burden on the affected populations as well as the healthcare system. Poor diagnosis may also contribute to the continued prevalence of the disease as many cases would remain undiagnosed and untreated, allowing the condition to persist and spread.

Studies conducted in Nigeria, Cameroon, and Tanzania have highlighted several factors contributing to the increased prevalence of FGS. These include socioeconomic disparities that influence the incidence of the disease among women, limited knowledge and awareness of FGS, poor healthcare-seeking behavior, poor access to treated water sources, lack of access to treatment, and healthcare support services(2,4,9). Other studies have highlighted additional contributing factors, such as a history of previous surgical procedures, HIV status, previous *S. haematobium* infection, and a lack of expertise among healthcare providers in delivering appropriate diagnosis and care(8,10,11). These factors contribute to the increased prevalence of the disease, posing challenges for effective interventions including integrated approaches.

There is a gap in Malawi regarding prevalence as well as the factors associated with FGS, as the area remains under-researched. Between 1974 and 1975, 43.5% (60/138) of women with infertility symptoms at the Zomba Hospital had schistosoma ova detected in their cervical biopsies(1). Furthermore, the histopathology of 176 cases of recognized gynecological schistosomiasis in Blantyre between 1976 and 1980 revealed that the cervix was affected in 60% of cases(1). In addition, a 1996 study revealed that genital biopsies revealed eggs in the cervix, vagina, and vulva in 65% (33/51) of women in the Mangochi district with S. haematobium in their urine(12). Lambert et al. (2024) showed that the prevalence of FGS in schistosomiasis-endemic areas in the Nsanje and Chikwawa districts was 26.9%. However, no studies have documented the burden of schistosomiasis in the endemic areas of central Malawi. This study aimed to assess the prevalence and risk factors of FGS among childbearing women in Nkhotakota District, Central Malawi.

## 2 Methods and Materials

### 2.1 Study Setting and Period

The study was conducted in public hospitals and health posts in Nkhotakota district, a lakeshore district, is situated in the central region of Malawi. Nkhotakota has a population of 463,771 with a total of 237,557 women. The population in the reproductive age group was 222,610, of which 106,667 were women. The district comprises 19 health centers, including dispensaries; four hospitals, including rural/community hospitals; and seven CHAM facilities. The study focused on three facilities: Nkhotakota District Hospital, a major referral hospital serving both urban and rural populations; Sasani Health Post, which caters to rural communities; and Sani Health Centre, a mid-level health facility positioned between a health post and a district hospital, serving predominantly rural populations. This study was conducted from November 2024 to March 2025.

### 2.2 Study Design and Population

This study employed a cross-sectional design, with multiple facilities. It focuses on a population of 102,906 women of childbearing age, specifically those aged between 18 and 49 years, residing in the Nkhotakota district. To be included, participants had to report symptoms or signs suggestive of genital schistosomiasis and provide consent to participate in the study. Women who were pregnant or menstruating at the time of data collection, or had a known history of cervical cancer, precancerous lesions, or prior cervical surgery were excluded from the study.

### 2.3 Sample Size Determination and Sampling Procedures

The sample size required for this study was calculated using the following prevalence study formula proposed by Pourhoseingholi(13):

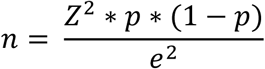

with the assumption of a 21.5% prevalence among school-going children according to the NTD program report(14). This resulted in a final sample size of 260 participants.

To obtain the sample, community sensitization was performed to encourage eligible women to attend a specified screening center. This method enabled engagement, but introduced the possibility of selection bias, especially self-selection bias. Participation was voluntary, meaning that women who attended may have systematically differed from those who did not. The data were collected from 260 participants.

### 2.4 Data Collection Methods and Procedure

A four-member team consisting of two clinical officers and two enumerators collected the data using an interviewer-guided questionnaire developed by the study team. The questionnaire had five sections: the first covered the socio-demographic characteristics of the participants, the second addressed knowledge and experience with schistosomiasis, the third focused on female genital schistosomiasis, the fourth assessed colposcopy feasibility and acceptance, and the final section presented findings from the FGS colposcopy screening. The data were collected between November 2024 and March 2025.

### 2.5 Variables

The primary outcome variable was binary-coded FGS status (*0 = negative, 1 = positive*), determined based on the professional judgment of a gynecologist following a colposcopic examination. Explanatory variables included sociodemographic characteristics, such as age (measured in years), residence (categorized as high- or low-burden village), occupation (e.g., farmer, entrepreneur, housewife, student, government employee, fishing), and level of education (ranging from no formal education to college level). Knowledge and experience of schistosomiasis were assessed through variables capturing prior awareness, history of infection, time since last infection, receipt of praziquantel treatment, and exposure to freshwater bodies, including bathing, washing clothes, farming, or swimming. Knowledge and experience with female genital schistosomiasis (FGS) were assessed through awareness, sources of information, and self-reported symptoms, such as genital itching, vaginal discharge, painful urination, blood in urine, pain during or bleeding after intercourse, lower abdominal pain, irregular or heavy menstruation, menstrual pain, difficulties conceiving, and forms of incontinence. Colposcopy findings included the presence of sandy patches in the mucosa, abnormal blood vessels, contact bleeding, edema, erosion, genital ulcers, malignant lesions, tumors, petechiae, and pain on bimanual palpation.

### 2.6 Ethical issues

The study was approved by the National Health Sciences Research Committee (NHRSC) in Malawi (approval number 24/03/4411). Participation in the study was voluntary, and all participants were assured of their privacy and confidentiality.

### 2.7 Data Processing and Analysis

Data were processed and analyzed using R (version 4.3.1) and the tidyverse suite of packages. Raw screening and behavioral/demographic data were cleaned, recoded, and merged using unique participant identifiers, resulting in a final analytical dataset comprising of 258 women with complete records. Data cleaning included standardization of participant IDs, relabeling of categorical variables, correction of inconsistencies, and exclusion of duplicate cases. Descriptive statistics were computed to summarize participant characteristics by FGS status, and associations were tested using chi-square or Fisher’s exact tests, as appropriate. A multivariable logistic regression model was constructed to identify predictors of FGS, with stepwise Archaic Selection Criteria (AIC)-based selection used to derive the most parsimonious model. Variance inflation factors confirmed no multicollinearity, and the model calibration was visually assessed. Results were reported as odds ratios (ORs) with 95% confidence intervals (CIs). All statistical analyses were conducted using a reproducible code with additional packages, including gtsummary, car, pROC, RMS, and resource selection.

## 3 Results

### 3.1 Baseline characteristics of the study population

Table 1 presents the baseline characteristics of the 258 study participants. The median age of the participants was 31 years (interquartile range: 25–38 years). The participants were nearly evenly distributed between high-burden (48%) and low-burden (52%) villages. Most participants were farmers (43%), followed by entrepreneurs (29%), and housewives (24%). More than half (56%) of the participants had not completed primary education, whereas only 1.2% had attained college-level education. A large proportion (93%) of participants had never heard of schistosomiasis. Among those with a history of schistosomiasis, 68% reported falling ill within the past 1–6 months and none reported receiving praziquantel treatment. Fifty-eight percent of the participants reported regular contact with freshwater bodies; among them, 33% had weekly exposure and 27% had contact more than three times per week. Additionally, 57% reported washing clothes, 71% bathing, and 76% swimming in water bodies. Only 27% of the participants reported having heard of FGS. Healthcare workers (54%) and friends/families (26%) were the most common sources of information.

**Table 1:** Baseline characteristics of study participants presented as frequencies and percentages for categorical variables and median and interquartile range for continuous variables

| Characteristic | Frequency | Percentage |
| --- | --- | --- |
| Age | 31(25,38) |  |
| Residence |  |  |
| High burden village | 123 | 48% |
| Low burden village | 135 | 52% |
| Occupation |  |  |
| Entrepreneur | 75 | 29% |
| Farmer | 112 | 43% |
| Fishing | 1 | 0.40% |
| Government employee | 5 | 1.90% |
| Housewife | 62 | 24% |
| No reply | 2 | 0.80% |
| Student | 1 | 0.40% |
| level of education |  |  |
| College | 3 | 1.20% |
| Completed primary education | 32 | 12% |
| Completed secondary education | 20 | 7.80% |
| No formal education | 21 | 8.10% |
| Not completed primary education | 144 | 56% |
| Not completed secondary education | 38 | 15% |
| heard schisto |  |  |
| No | 239 | 93% |
| Yes | 19 | 7.40% |
| suffered schisto |  |  |
| No | 107 | 41% |
| Yes | 151 | 59% |
| length of illness |  |  |
| >24 months | 4 | 3.70% |
| 1-6 months | 73 | 68% |
| 12 to 24 months | 11 | 10% |
| 6-12 months | 19 | 18% |
| Praziquantel |  |  |
| No | 258 | 100% |
| contact freshwater |  |  |
| No | 113 | 44% |
| Yes | 145 | 56% |
| water contact freq |  |  |
| More than three times a week | 31 | 27% |
| Once a week | 37 | 33% |
| Once in 2 weeks | 16 | 14% |
| Two to three times a week | 29 | 26% |
| wash waterbodies |  |  |
| No | 111 | 43% |
| Yes | 147 | 57% |
| bath waterbodies |  |  |
| No | 76 | 29% |
| Yes | 182 | 71% |
| swim waterbodies |  |  |
| No | 63 | 24% |
| Yes | 195 | 76% |
| heard female gen schisto |  |  |
| No | 189 | 73% |
| Yes | 69 | 27% |
| source of information |  |  |
| Community leaders | 1 | 0.50% |
| Friends/family | 50 | 26% |
| Healthcare worker | 103 | 54% |
| Media (radio, TV, posters) | 33 | 17% |
| School | 2 | 1.10% |

### 3.2 Self-reported signs and symptoms by FGS status

The study participants self-reported the following signs and symptoms: genital itching, vaginal discharge, pain when urinating, blood in the urine, pain during and after sex, bleeding after sex, lower abdominal pain, spot bleeding, irregular menstruation, heavy bleeding, menstrual pain, difficulty getting pregnant, stress incontinence, urge incontinence, anemia, and menstrual period for three weeks or more. Blood in urine and bleeding after sex were negatively associated with FGS status, with both signs showing a higher prevalence in women who were FGS-negative than in women who were FGS-positive (Table 2).

**Table 2:**
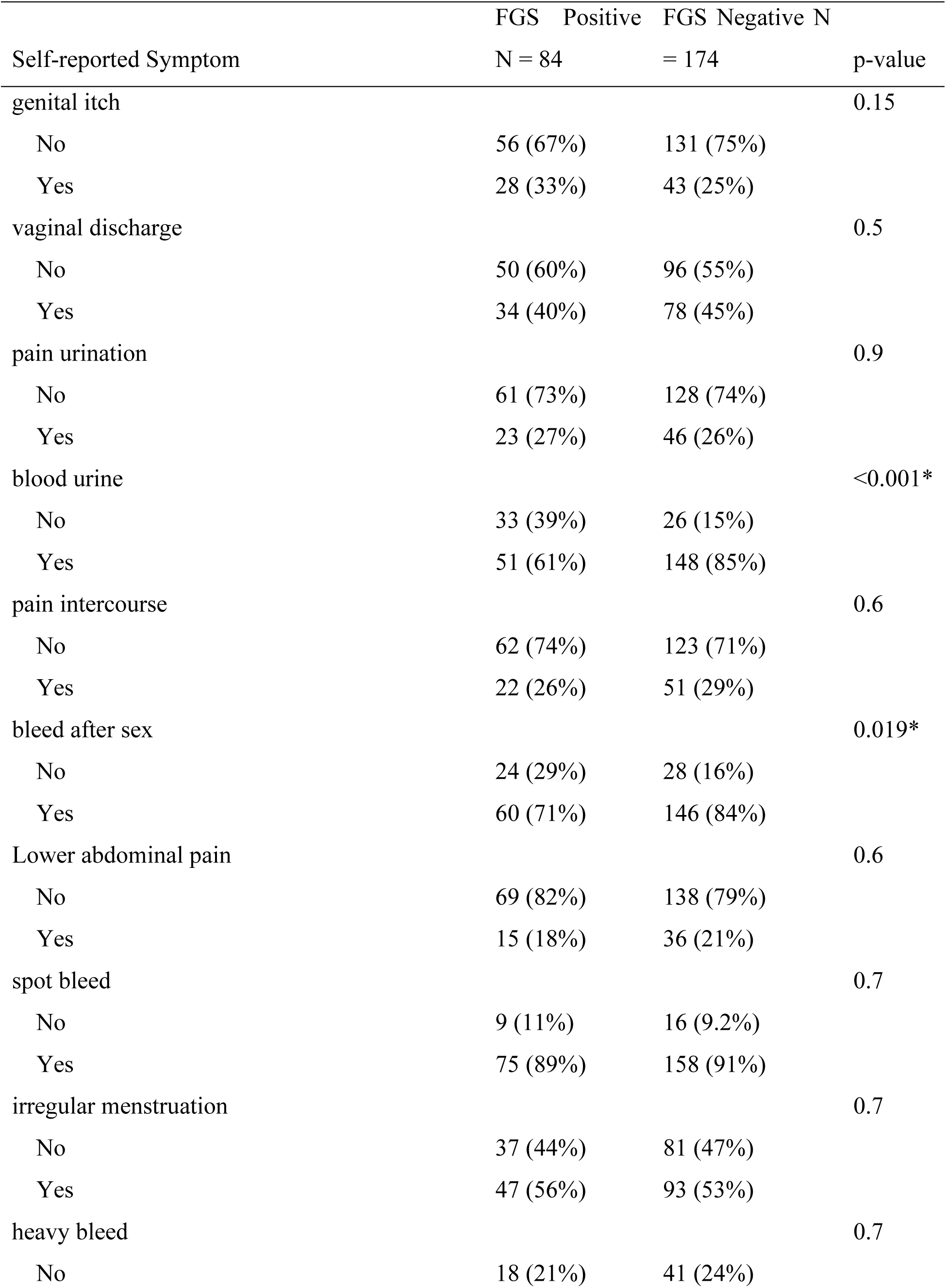
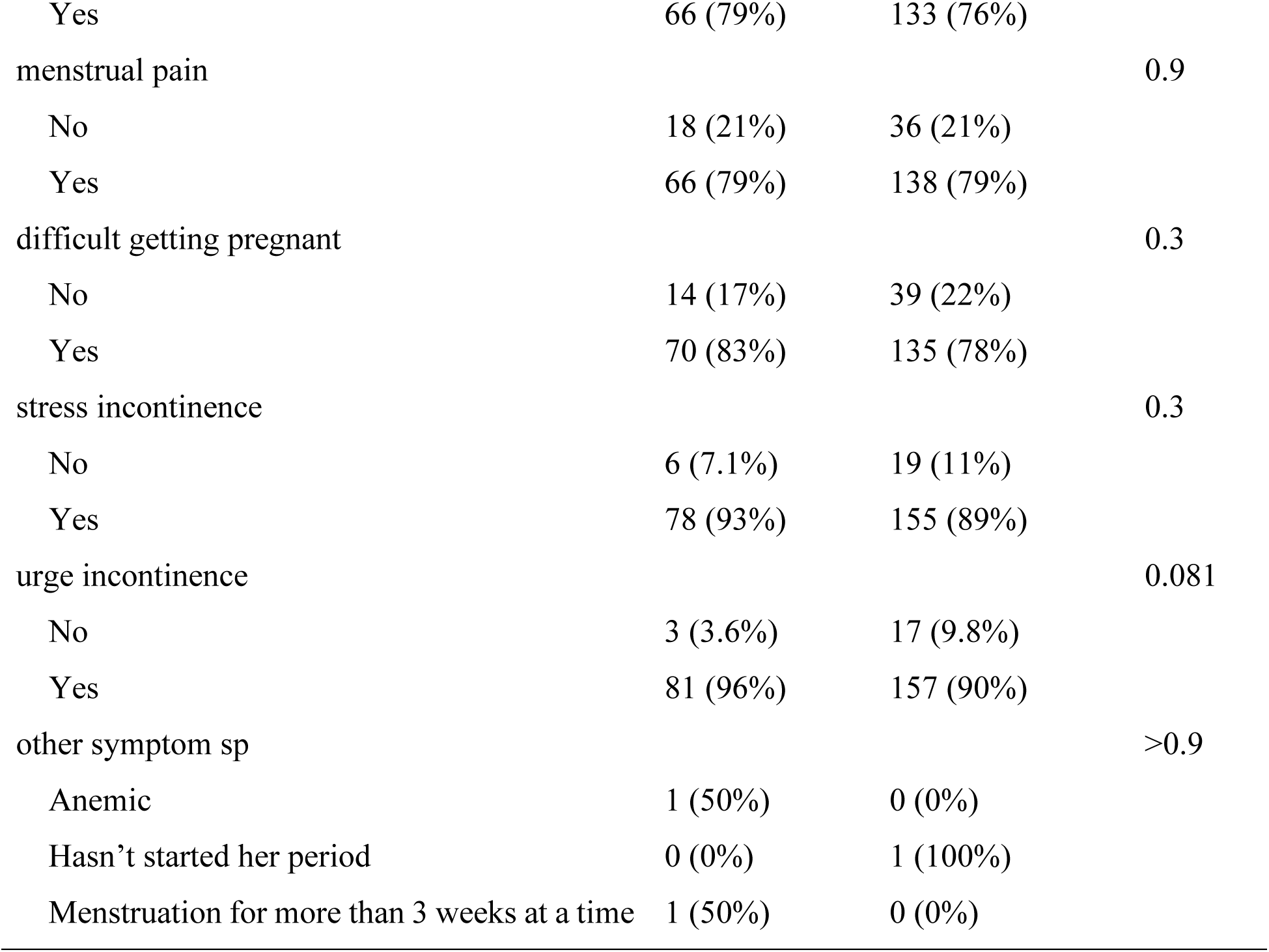
Association between self-reported symptoms and female genital schistosomiasis Status

### 3.3 Prevalence of FGS and the distribution of colposcopy lesions by FGS status

Table 3 presents the prevalence of FGS and the distribution of lesions found on colposcopy according to the FGS status. The prevalence of FGS based on colposcopy across the study population was 32.6% (84/258). Several colposcopy findings, except for cervical edema, genital ulcer, and pain on bimanual palpation, were significantly associated with FGS positivity, indicating strong diagnostic relevance (Table 3). Abnormal blood vessels and sandy patches in the mucosa were the most common lesions found during colposcopy, with a prevalence of 69% and 68%, respectively, among the FGS-positive participants. The other lesions were found in varying proportions (Table 3).

**Table 3:** Colposcopy findings by FGS status among study participants

| **Variable** | **Overall N = 258** | **Positive N = 84 (32.6%)** | **Negative N = 174 (67.4%)** | **p-value** |
| --- | --- | --- | --- | --- |
| *Overall Prevalence* | *84(32.6%)* |  |  |  |
| Sandy patches in mucosa |  |  |  | <0.001 |
| no | 195 (76%) | 27 (32%) | 168 (97%) |  |
| yes | 63 (24%) | 57 (68%) | 6 (3.4%) |  |
| Abnormal blood vessel |  |  |  | <0.001 |
| no | 189 (73%) | 26 (31%) | 163 (94%) |  |
| yes | 69 (27%) | 58 (69%) | 11 (6.3%) |  |
| Contact bleeding |  |  |  | 0.003 |
| no | 237 (92%) | 71 (85%) | 166 (95%) |  |
| yes | 21 (8.1%) | 13 (15%) | 8 (4.6%) |  |
| edema |  |  |  | 0.2 |
| no | 212 (82%) | 65 (77%) | 147 (84%) |  |
| yes | 46 (18%) | 19 (23%) | 27 (16%) |  |
| erosion |  |  |  | <0.001 |
| no | 241 (93%) | 71 (85%) | 170 (98%) |  |
| yes | 17 (6.6%) | 13 (15%) | 4 (2.3%) |  |
| genital ulcer |  |  |  | 0.11 |
| no | 256 (99%) | 82 (98%) | 174 (100%) |  |
| yes | 2 (0.8%) | 2 (2.4%) | 0 (0%) |  |
| Malignant lesion |  |  |  | 0.002 |
| no | 250 (97%) | 77 (92%) | 173 (99%) |  |
| yes | 8 (3.1%) | 7 (8.3%) | 1 (0.6%) |  |
| genital tumor |  |  |  |  |
| no | 258 (100%) | 84 (100%) | 174 (100%) |  |
| petechiae |  |  |  | <0.001 |
| no | 230 (89%) | 63 (75%) | 167 (96%) |  |
| yes | 28 (11%) | 21 (25%) | 7 (4.0%) |  |
| pain bimanual palpation |  |  |  | 0.09 |
| no | 225 (87%) | 69 (82%) | 156 (90%) |  |
| yes | 33 (13%) | 15 (18%) | 18 (10%) |  |

### 3.4 Risk factors associated with FGS

Multivariate logistic regression analysis was conducted to evaluate possible risk factors associated with FGS (Table 4). Age, self-reported symptoms such as blood in the urine, blood after sex, and previous cervical cancer screening were significantly associated with the FGS status. Increasing age increased the odds of FGS, with each additional year linked to a 5% increase in odds (OR, 1.05; 95% CI: 1.02–1.09; p = 0.003). Similarly, having reported previous cervical cancer screening was associated with increased odds of FGS compared to those who did not report having previously undergone cervical cancer screening (OR: 1.91; 95% CI: 1.01– 3.70; p = 0.05), and interestingly, individuals who reported contact with freshwater had significantly lower odds of FGS compared to those who had no contact with fresh water (OR: 0.55; 95% CI: 0.30–0.98; p = 0.045). Those who self-reported blood in urine were less likely to have FGS than those who did not report blood in urine (OR: 0.25; 95% CI: 0.13–0.49; p < 0.001). Similarly, those who reported bleeding after sexual intercourse were also less likely to have FGS than those who did not report bleeding (OR, 0.43; 95% CI: 0.21–0.85; p = 0.016). Other variables, including awareness of FGS, genital itching, pain during urination, and urge incontinence, did not show statistically significant associations, although some trends suggested possible relationships, warranting further investigation.

**Table 4:**
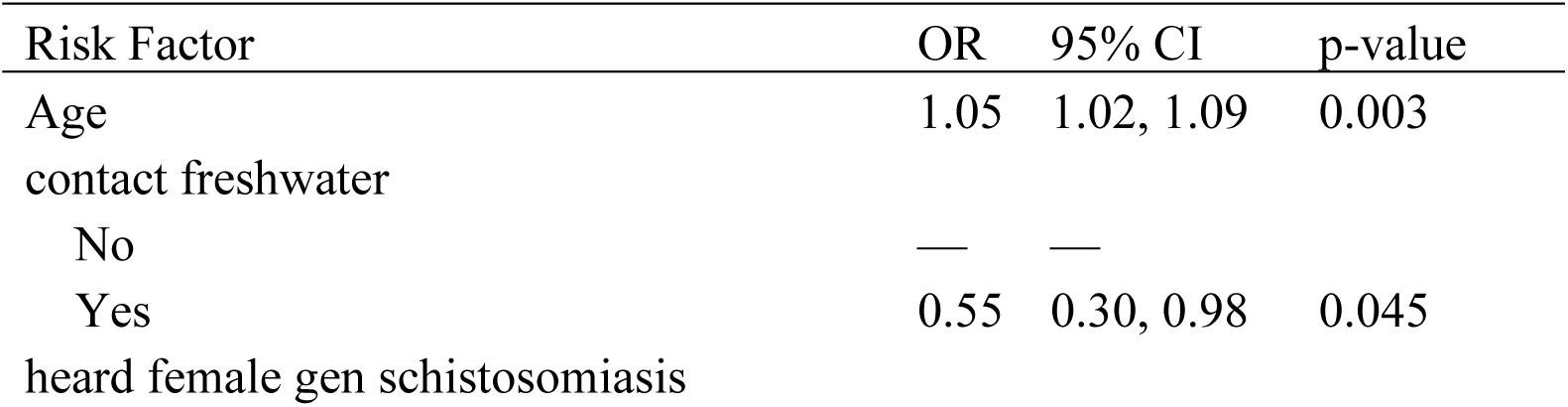
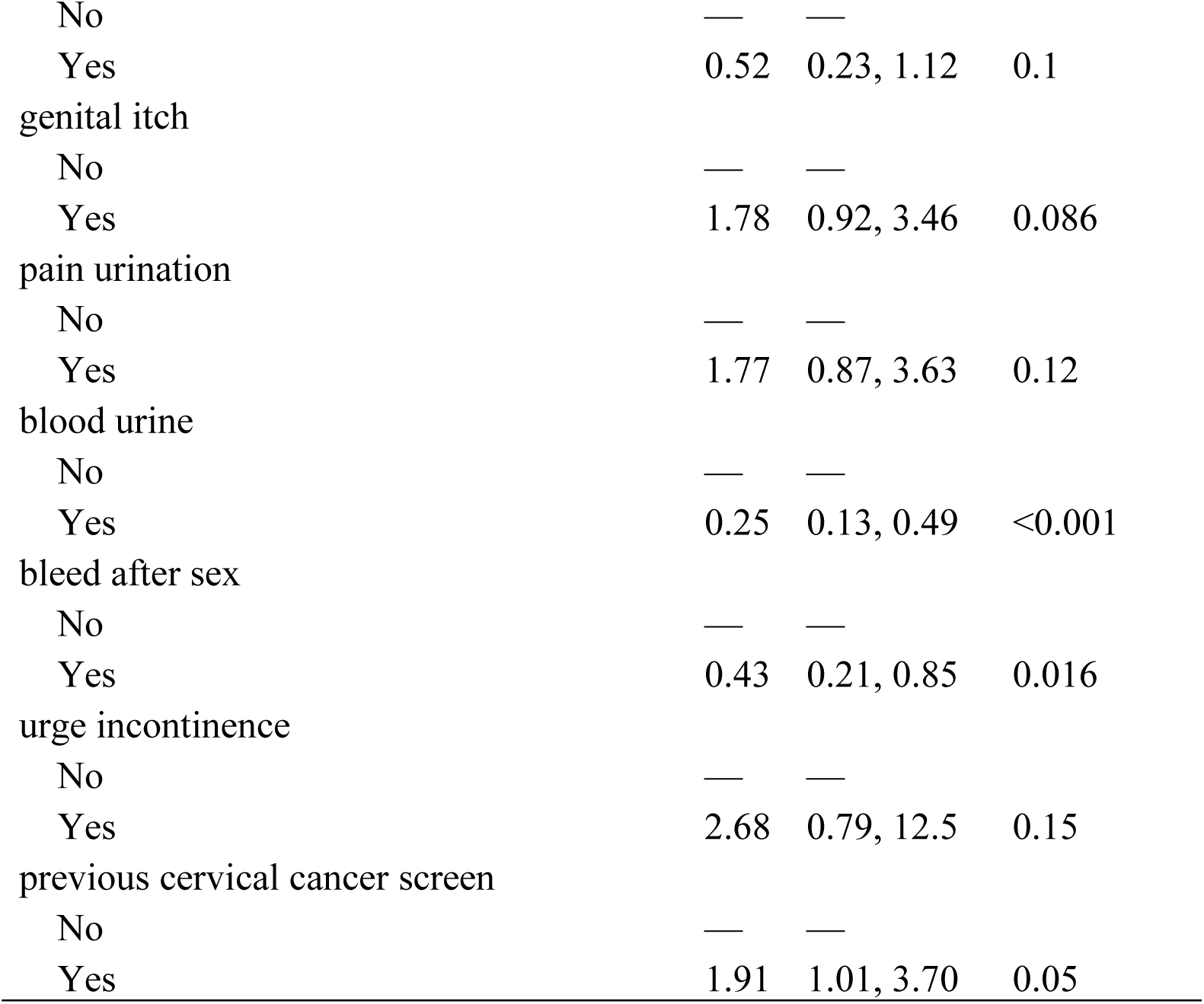
Multivariable logistic regression of risk factors associated with female genital schistosomiasis

#### 3.4.1 Validation of Risk Factors for Clinical Prediction Modelling of FGS using Observed Data

The predictive performance of the final model was assessed using calibration and discrimination metrics. The calibration plot (Figure 1) demonstrated excellent agreement between the predicted probabilities and observed event rates, with a near-perfect alignment along the diagonal (mean absolute error = 0.001), indicating that the probabilistic predictions of the model were well-calibrated to the ground true frequencies. The receiver operating characteristic (ROC) curve (Figure 2) further evaluated the model’s classification capability across decision thresholds, showing robust sensitivity-specificity tradeoffs. While the full specificity range is not displayed in the available data, the trajectory of the curve suggests strong discriminatory power, with sensitivity values approaching the ideal range (0.0–1.0) for practical classification tasks. The combination of low calibration error and high discriminative performance implies that the model is both statistically reliable and clinically (or experimentally) actionable for its intended use. Further quantification of the area under the ROC curve (AUC) would provide additional validation of the classification efficacy.

**Figure 1:**
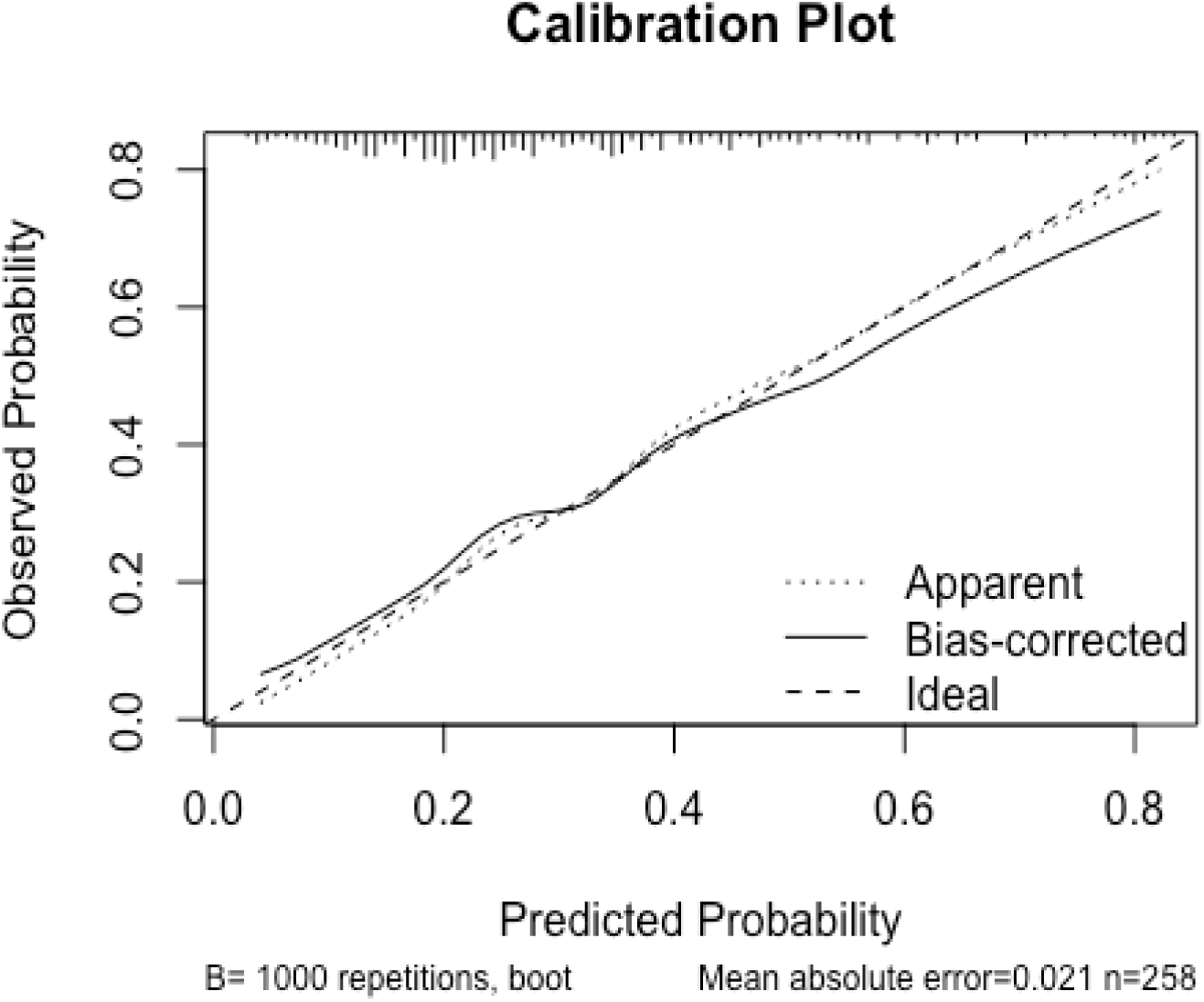
1 Calibration performance of the predictive model. The diagonal dashed line represents perfect calibration (predicted probabilities = observed frequencies). Points show the model’s predicted probabilities (x-axis) versus empirical event rates (y-axis) across the probability bins. The reported mean absolute error (MAE = 0.001) indicates near-ideal calibration. Error bands (not shown) would typically represent 95% confidence intervals from 1,000 bootstrap repetitions

**Figure 2:**
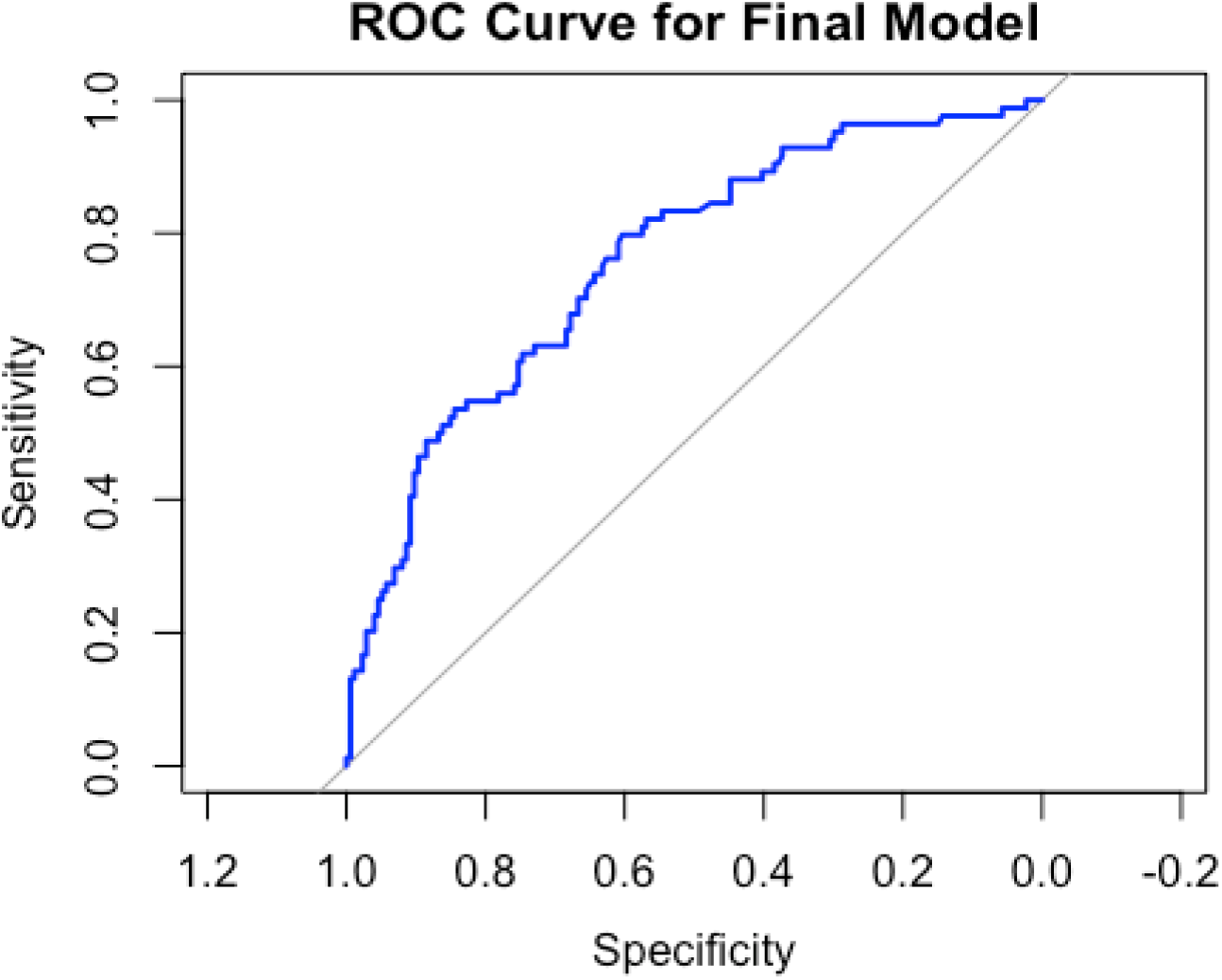
Receiver Operating Characteristic (ROC) curve for the final model. Sensitivity (true positive rate) is plotted against 1 – specificity (false positive rate) across the classification thresholds. The dashed diagonal line represents the random chance (AUC = 0.5). The upward-concave trajectory suggests discriminative power exceeding chance, although the full specificity values are truncated. The area Under the Curve (AUC) and 95% confidence intervals are reported in the figure or accompanying text.

## 4 Discussion

This study investigated the prevalence of and factors associated with FGS in the Nkhotakota district among women of childbearing age using the visual diagnostic method of colposcopy. To our knowledge, this is the first study to assess the burden of FGS in the central region of Malawi. Our study demonstrated a high FGS burden in the Nkhotakota district. In terms of factors associated with FGS, we found a statistically significant association between age and self-reported symptoms, such as blood in the urine and blood after sex, and having previously undergone cervical cancer screening with FGS status. We found no statistically significant associations between self-reported renal, sexual, and reproductive health (SRH) symptoms and FGS status.

We found the prevalence of FGS to be 32.6%, which was higher than the prevalence of visual FGS in southern Malawi (26.9%)(15). Other studies from other countries have also reported a lower prevalence, ranging from 4% in Egypt to 23% in South Africa(16–20). In contrast, other studies conducted in Zambia, Ghana, Tanzania, Cameroon, and Nigeria found a higher prevalence of FGS among women, ranging from 37% to 79.5%(21–24). The high burden of FGS in the current study suggests that women are at an increased risk of developing FGS-associated health problems, including infertility and sexually transmitted diseases, such as HIV. Previous studies have shown that women with FGS are four times more likely to acquire HIV, which is attributed to bleeding during sex(10,25). This stresses the need to avoid sexual intercourse when diagnosed with FGS, until it is treated. Other studies have recommended the integration of FGS in sexual reproductive health programs, as it may increase the chances of contracting sexually transmitted infections among those involved in sexual activity. This may be a better strategy to enhance our understanding of this condition and its effects(6,26).

In the current study, sandy patches were the second most prevalent lesions after abnormal blood vessels in FGS-positive women. This is supported by previous evidence that sandy patches are the hallmark features of FGS(2,27). These are caused by persistent inflammation due to schistosome eggs stuck in the tissues of the pelvic organs, bladder, lower uterus, cervix, vagina, prostate gland, and seminal vesicles, which contributes to the development of sandy patches(4). The presence of these sandy patches may inform healthcare workers to provide the right treatment, as they are part of the signs of FGS. The study further found that abnormal blood vessels were the most prevalent lesions among FGS-positive women. Other studies agree that the presence of abnormal blood vessels on the cervical surface indicates a positive case of FGS(2,27,28). Contact bleeding was also prevalent among women who were FGS-positive. This refers to bleeding that occurs upon physical examination or contact with the genital mucosa, which is not attributable to menstruation or trauma. Contact bleeding has also been reported in other studies, showing a relationship between contact bleeding and FGS(2,29). These signs may inform health workers to easily make a diagnosis and provide necessary support to women.

In terms of risk factors for FGS, the current study found a significant relationship between age and FGS development. Evidently, an increase in age places additional household responsibility. In Malawi, as girls mature, they are involved in various household activities, such as water fetching, washing clothes, farming, and bathing younger children. While undertaking these roles, women may end up using contaminated water, increasing their chances of contracting FGS. Nkhotakota district is a lakeshore area where women frequently visit the lake and use water from the lake for various purposes. This exposes them to the risk of acquiring FGS.

Regarding water contact, this study found that individuals who reported contact with freshwater had significantly lower odds of having FGS. This finding contradicts the general understanding that exposure to water sources is a risk factor for FGS(1,30). Individuals who frequently come into contact with various water sources are health-conscious and, therefore, more likely to seek treatment when FGS symptoms are observed. It is also likely that some freshwater contact points are not infested by intermediate host snails, which are part of the schistosome life cycle. However, it is important for women to take appropriate measures when it comes to the use of water from the lake, as people from lakeshore areas depend on the lake for their daily living activities such as fishing, bathing, and farming. Previous studies in Malawi have also noted that some people in lakeshore areas do not have toilets because they relieve themselves from the lake, which makes it more contaminated (31,32).

This study assessed the relationship between previous cervical cancer screening and development or detection of FGS. Participants who underwent cervical cancer screening had nearly twice the odds of having an FGS. Women who visit hospitals for cervical cancer may undergo comprehensive vaginal examination, which may also result in the detection of symptoms related to FGS. Furthermore, women who may experience pain and other vaginal discharges may opt for cervical cancer screening when FGS is detected. Healthcare seeking behavior among women seeking cervical cancer screening may result in the detection of FGS.

The findings of this study indicate a strong association between bleeding after sexual intercourse and the presence of FGS. This is caused by chronic inflammation and damage caused by Schistosoma haematobium eggs deposited in the genital tissues. These eggs induce reactions and tissue fibrosis, leading to friable mucosal surfaces that are prone to bleeding during sexual activity. Furthermore, the fragile genital mucosa and vaginal wall become susceptible to intercourse injury, resulting in bleeding(2,4,9). Genital bleeding may have significant effects on women in terms of health, psychosocial, and reproductive health implications, including stigma, sexual dysfunction, and relationship issues(33). This requires the implementation of awareness programs among women. Once they observe bleeding, they should seek healthcare support. Women who may have such experiences must seek timely treatment from healthcare facilities to address this problem.

The results of this study were validated and demonstrated the discriminative and calibrated nature of the FGS prediction model. The nearly flawless alignment of the calibration plot (mean absolute error = 0.001) shows that the expected probabilities and actual results were similar. Although the entire specificity range and AUC were not disclosed, the ROC curve indicated good sensitivity and classification performance. These results collectively bolster the model’s dependability and possible clinical utility in identifying people at risk for FGS.

## Implications and recommendations

To lessen the burden of FGS in endemic populations, such as lakeshore areas, these findings recommend greater water sanitation initiatives and increased public knowledge of early warning indicators of FGS to seek timely healthcare interventions. There is also a need for the integration of FGS into reproductive health screening and awareness programs to enhance the early recognition of the disease and enhance the understanding of its severity and transmission dynamics in the general population. The high burden of FGS among women may result in an increased risk of schistosomiasis infection among children under five years of age, who most likely accompany their mothers to contaminated freshwater bodies. To reduce the risk in this age group, health authorities should explore the introduction of an integrated pediatric praziquantel program for children under five years. For a comprehensive and integrated approach, health authorities should undertake snail surveys at freshwater contact points to establish the presence or absence of intermediate host snails, and undertake appropriate control and community awareness measures.

## Strengths and limitations

Our study has some limitations. We measured the presence of FGS using visual colposcopy, which is not the gold standard for definitive diagnosis of schistosome lesions. This may have led to overestimation of the burden in this study. Its cross-sectional nature means that causality between an FGS positive status and risk factors cannot be established. While the study was powered to estimate the burden of FGS in Nkhotakota District, it may not necessarily reflect the situation in other areas of Malawi. The study did not assess the participants’ knowledge of the association between schistosomiasis and intermediate host snails, an additional risk factor that would influence their decision to avoid specific freshwater contact points or activities.

Despite these limitations, this study had several strengths. To the best of our knowledge, this is the first study to characterize FGS in Central Malawi. The sample size used in this study met the minimum required to estimate the burden of FGS in the study area. Moreover, the sampling method used in this study minimized the possibility of selection bias, with positive implications for the study findings.

## Conclusion

There is a huge burden of FGS among women of childbearing age in the Nkhotakota District, central Malawi. The factors associated with FGS include increasing age and a history of cervical cancer screening. Contextualized interventions directed at preventing and controlling FGS among women of childbearing age should be identified and implemented in schistosomiasis endemic areas. Authorities should consider integrating FGS screening with existing cervical cancer screening clinics and/or mother-child health services for early diagnosis and treatment. Larger studies are required to estimate FGS burden at the national level. In addition, further studies are needed to investigate the feasibility of integrating FGS screening services into existing services, such as cervical cancer screening. Future research should assess the readiness of countries to introduce pediatric praziquantel programs as part of schistosomiasis prevention efforts.

## Declarations

### Human Ethics and Consent to Participate declarations

Applicable. Ethical approval was granted by the National Health Sciences Research Committee (NHSRC), Malawi (Approval number: 24/03/4411), and all participants provided written informed consent.

### Consent for publication

Not applicable

### Availability of data and study resources

The data for this study are available from the lead author upon request.

### Authors’ contributions (initials)

All authors meet the criteria for authorship as stated in the International Committee of Medical Journal Editors (ICMJE) authorship guidelines. DMK, ECB, AT, CS, and EMK conceived of and designed the study. DMK, ECB, and MK secured the grant funding. AT, DMK, and MK analyzed the data. AT and MK designed the online dashboard for data collection. DMK, CS, EMK, FM, TM, JF, and CM provided the methodological insights. AT, MK, and DMK oversaw the data curation. CM, SL, TM, and FM collected data. DMK, ECB, EMK, AT, CS, JM and MK drafted the manuscript. ECB, MK, DMK, EMK, and MK supervised this study. All the authors reviewed and commented on the manuscript and approved the final draft for publication.

## Data Availability

Data used in this submission can be accessed via the provided link https://1drv.ms/x/c/9a64aedb6dfb7c1b/EeWBoexrUzdOlDMeFqQtaxsBa3UTU3Tyitlg4JtXmFq1JQ

## Acknowledgements

We acknowledge the participants for their time spent answering our questions and consenting to the screening procedure.

